# A mixture of essential oils from three Cretan Aromatic Plants (thyme, Greek sage and Cretan dittany, CAPeo) inhibits SASR-CoV-2 proliferation: *in vitro* evidence and a Proof-of-Concept intervention study in mild ambulatory COVID-19-positive patients

**DOI:** 10.1101/2021.01.11.20248947

**Authors:** Christos Lionis, Ioannis Karakasiliotis, Elena Petelos, Manolis Linardakis, Athanasios Diamantakis, Emmanouil Symvoulakis, Maria Panopoulou, Marilena Kampa, Stergios A. Pirintsos, George Sourvinos, Elias Castanas

**Affiliations:** Clinic of Social and Family Medicine, School of Medicine, University of Crete, Heraklion, Greece; Laboratory of Biology, School of Medicine, Democritus University of Thrace, Alexandroupolis, Greece; Laboratory of Clinical Microbiology, University General Hospital, Alexandroupolis, Greece; Laboratory of Microbiology, Department of Medicine, Democritus University of Thrace, Alexandroupolis, Greece; Laboratory of Experimental Endocrinology, School of Medicine, University of Crete, University of Crete, Heraklion, Greece; Department of Biology, School of Sciences and Technology, University of Crete, Heraklion, Greece; Botanical Garden, University of Crete, Rethymnon, Greece; Laboratory of Clinical Virology, School of Medicine, University of Crete, Heraklion, Greece

**Keywords:** SARS-CoV-2, COVID-19, natural products, essential oils, therapeutic agent

## Abstract

The need for therapeutic regimens for the non-critically ill patients of the COVID-19 pandemic remains unmet. In this line, repurposing existing drugs, against known or predicted SARS-CoV-2 protein actions, has been advanced, while natural products have also been tested. Previous work has shown that a Cretan Aromatic Plant (*Thymbra capitata* (L.) Cav., *Salvia fruticosa* Mill. and *Origanum dictamnus* L.) essential oil mixture (CAPeo) has a remarkable *in vitro* antiviral activity against Influenza A & B and Rhinovirus 14 strains, decreasing the symptoms of upper respiratory tract infections, while proven safe in experimental animals and humans. Here, we tested CAPeo in VERO cells infected with SASR-CoV-2. We report that this mixture, at similar concentrations as those previously reported, exhibits a remarkable antiviral activity. Administration of 1 ml of a 1.5% CAPeo in olive oil, in a Proof-of-Concept intervention study in SARS-CoV-2-positive, exhibiting mild COVID-19 symptoms, humans resulted in a significant amelioration of general and local symptoms of the disease. We conclude that CAPeo may be a valuable addition for the prevention and/or treatment of mild COVID-19 ambulatory patients, pending a confirmation through a prospective randomized controlled trial in humans (NCT04705753).

## Introduction

Since the outbreak of COVID-19 pandemics, in 2019, a previously unseen international effort has been undertaken for the identification of the underlying cause (the SARS-CoV-2 virus) and the detailed analysis of its genome (O’Leary and Ovsepian, 2020). An international effort is actually directed towards an efficient therapy (Baum et al., 2020;Hansen et al., 2020;Weinreich et al., 2020), or the development of efficient vaccines (Polack et al., 2020;Voysey et al., 2020). A number of established pharmaceutical molecules and patients’ plasma have been tested as drug candidates for the treatment of COVID-19, with variable results (see (Bolarin et al., 2020;Wang et al., 2020a;Wang et al., 2020b) and references herein). Among the multitude of products tested against COVID-19 disease, a number of natural products, including herbal extracts, have also been assayed (critically reviewed in (Benarba and Pandiella, 2020) and references herein), targeting mainly the viral proteases.

Recently, we have reported that a combination of three aromatic plants essential oil (CAPeo) (*Thymbra capitata (L*.*) Cav*., *Origanum dictamnus L*., *Salvia fruticosa Mill*., (Pirintsos et al., 2020), and references herein) is efficient against upper respiratory tract viral infections, in humans (Duijker et al., 2015;Anastasaki et al., 2017). *In vitro* studies revealed the efficacy of CAPeo against Influenza A & B and Human Rhinovirus 14, and reported an action through the inhibition of the nuclear translocation of viral nucleoproteins (Tseliou et al., 2019), resulting in impaired viral protein transcription. Furthermore, we have reported the safety of CAPeo, both in humans (administered in the form of soft gels, 1 ml/day of a 1.5% essential oil combination in extra virgin olive oil, (Duijker et al., 2015)) and in experimental animals (Kalyvianaki et al., 2020). In the present study, we have assayed the efficiency of CAPeo mixture on the proliferation of SARS-CoV-2 in VERO cells. We report a remarkable antiviral activity of CAPeo, at concentrations compatible with those obtained after the recommended dose administration in humans. Moreover, we performed a Proof-of-Concept intervention study in mild COVID-19-positive humans and report that CAPeo can significantly ameliorate the general and local symptoms of the disease. We suggest that CAPeo, pending additional confirmation of results through a prospective randomized controlled trial, may represent a valuable addition for the prevention and/or therapeutic management of mild COVID-19 ambulatory patients.

## Material and Methods

### CAPeo production and use

Spanish oregano (*Coridothymus capitatus* (L) Rchb. F. synonym of *Thymbra capitata* (L) Cav.), dictamnus or Cretan dittany (*Origanum dictamnus* L) and sage (*Salvia fruticosa* Mill., *Salvia pomifera* L., were cultivated under total Good Agricultural Practice and high precision agriculture, based on an Ecological Niche Modelling tool, we have recently developed (Bariotakis et al., 2019), in order to maximize their essential oil composition and content. A constant genotype of plants, specified by a barcoding of each batch, was used. Essential oils were prepared by steam distillation of the dried plant leaves, under GMP conditions. The final extract, contained 4 parts *Corydothymus Capitatus* (L) extract, 2 parts *Salvia Fruticosa* Mill. extract and 1 part *Origanum Dictsmnus* L extract. It was analyzed by Gas Chromatography–Mass Spectroscopy (GC–MS), in a Shimadzu, QP 5050A apparatus. The mixture of essential oils contains carvacrol (53%) eucalyptol (13%) and β-Caryophyllene (3%). Concentrations of the compounds p-Cymene, γ-Terpinene, Borneol and α-Terpineol were 1.32, 1.17, 1.68 and 1.06% respectively, while the concentrations of the remaining 15 compounds were less than 1%. For the complete analysis of compounds, please refer to previous reports (Duijker et al., 2015;Kalyvianaki et al., 2020). These concentrations refer to the stock essential oil mixture, while a concentration of 1.5% in DMSO (Sigma-Aldrich) was used in the present study. This refers to the dilution 1/1, mimicking the suggested daily dose of the CAPeo extract in humans (1 ml of a 1.5% of CAPeo in olive oil, for the management of upper respiratory tract infections (Duijker et al., 2015;Anastasaki et al., 2017)). As the pharmacokinetics and bioavailability of CAPeo are under investigation, we have used bibliography data, suggesting a variable absorption of phenolic compounds ranging from 27 to 0.0006% and a blood recovery ≤1% for the majority of compounds (Scalbert et al., 2002;Manach et al., 2005). Therefore, to mimic available concentrations in humans, different dilutions (1:10, 1:100 and 1:1000 of the clinically administered concentration -15 mL extract/L, 1 mL/day-) in DMSO) were used in the present study. The same concentrations were used in a previous study, to determine the protective and therapeutic effect of CAPeo in cells infected with other upper respiratory viruses (Tseliou et al., 2019).

### Virus and virus titration

SARS -CoV-2 (isolate 30-287) was obtained through culture in Vero E6 cells, from an infected patient, in Alexandroupolis, Greece. Virus stock was prepared by infecting fully confluent Vero E6 cells in DMEM, 10% fetal bovine serum (FBS), with antibiotics at 37°C, 5% CO_2_. Four days after inoculation, the supernatant was frozen at −80°C until use. Titration was carried-out in 96 -well plates using Vero E6 cells and TCID_50_ was calculated according to the method of Reed and Muench (Reed and Muench, 1938). Plates were incubated at 37°C for 4 days, and the cytopathic effect (CPE) was scored by observation under an inverted phase contrast microscope.

### Infections and Treatment

Infections were carried out in 96-well plates, using SARS -CoV-2 (m.o.i. of 0.1) on Vero E6 cells. Cells were treated with different concentrations of CAPeo, as described above, in a volume of 15 μl, per 150 μl of medium, for 48h. Cell morphology was observed with phase contrast, in an inverted microscope, to record CPE. Culture supernatants were also collected and analyzed using real-time RT-PCR.

### Real-time RT-PCR

To determine viral load, RNA was extracted from 96-well supernatants (100 μl) using NucleoSpin Dx Virus according to the manufacturer (Macherey Nagel). Multi-target real-time RT-PCR was performed using COVID-19 SARS-Cov-2 Real-TM according to the manufacturer (Sacace Biotechnologies, Como, Italy).

### Patients included in the study

Seventeen (17) young adult patients (34.4±11 years) were included in a Proof-of-Concept intervention study, reporting to a single primary care unit with symptoms, related to an upper respiratory tract infection. SARS-CoV-2 infection was confirmed by real-time PCR, performed in the regional COVID-19 reference centre for COVID infections (Laboratory of Clinical Virology, University of Crete, School of Medicine). The study was conducted from Sep 1 to Oct 15, 2020. In parallel to the eligible patients, data for two family members (real-time PCR negative) of one infected participant, were included in the study analysis (see Results). The CAPeo mixture, in the form of two 0.5 ml soft capsules, in a concentration of 15 ml/L, was administered daily for two weeks (14 days), *per os*. Information regarding demographics, medical history, smoking habits, symptoms and signs has been recorded in a pre-tested questionnaire for all study participants (Table 1).

**Table 1.**
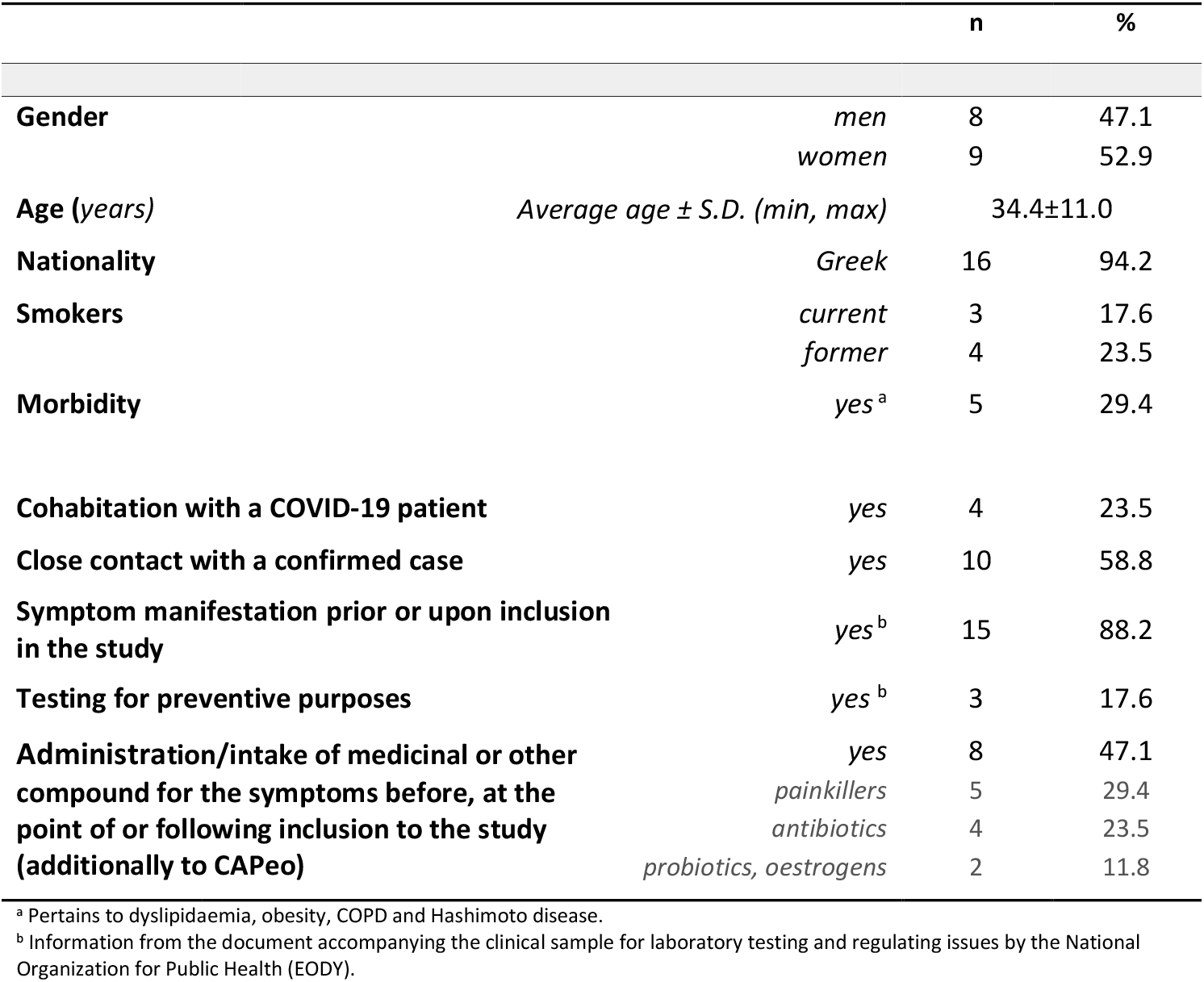
Descriptive characteristics of the 17 patients included in the study

|  |  | n | % |
| --- | --- | --- | --- |
| **Gender** | *men* | 8 | 47.1 |
|  | *women* | 9 | 52.9 |
| **Age (years)** | *Average age ± S.D. (min, max)* | 34.4±11.0 |  |
| **Nationality** | *Greek* | 16 | 94.2 |
| **Smokers** | *current* | 3 | 17.6 |
|  | *former* | 4 | 23.5 |
| **Morbidity** | *yes<sup>a</sup>* | 5 | 29.4 |
| **Cohabitation with a COVID-19 patient** | *yes* | 4 | 23.5 |
| **Close contact with a confirmed case** | *yes* | 10 | 58.8 |
| **Symptom manifestation prior or upon inclusion in the study** | *yes<sup>b</sup>* | 15 | 88.2 |
| **Testing for preventive purposes** | *yes<sup>b</sup>* | 3 | 17.6 |
| **Administration/intake of medicinal or other compound for the symptoms before, at the point of or following inclusion to the study (additionally to CAPeo)** | *yes* | 8 | 47.1 |
|  | *painkillers* | 5 | 29.4 |
|  | *antibiotics* | 4 | 23.5 |
|  | *probiotics, oestrogens* | 2 | 11.8 |

<sup>a</sup> Pertains to dyslipidaemia, obesity, COPD and Hashimoto disease.
<sup>b</sup> Information from the document accompanying the clinical sample for laboratory testing and regulating issues by the National Organization for Public Health (EODY).

Data were collected on day 1, day 4, day 7 and day 14; following the initial face-to-face consultation, data collection and consultations were performed, either remotely (by phone), or via home visits, by trained medical personnel. The severity of symptoms was assessed through the utilisation of a seven-point Likert scale, with data recorded on Day 1 and taken as the baseline. A seven-point Likert scale allowed recording of reported symptoms starting from 1 (minor) to 2 (very mild) and 3 (mild), 4 (somewhat moderate) and 5 (moderate) and culminating to 6 (severe) and 7 (very severe). The main outcome defined to assess the clinical effectiveness of the CAPeo was symptom reduction, in terms of severity and frequency, defined as total number of symptoms over the 14-day period, and with measurement on day 4, day 7 and day 14. Five patients reported symptoms prior to their confirmed diagnosis; the corresponding interval varied from 2 to 4 days. For the purposes of reporting and given the small interval prior to the confirmed diagnosis and study inclusion, the date of the confirmed diagnosis, i.e., day 1 of the study was considered as day 1 for symptoms, and severity and frequency assessment.

### Statistical analysis

Data were analyzed using the SPSS software (IBM SPSS Statistics for Windows, Version 26.0 Armonk, NY: IBM Corp) and Origin Pro 2018 (Originlab Co, Nothampton, MA). A critical value of 0.05 was taken as the threshold of statistical significance.

### Ethics

The study received approval by the University of Crete Bioethics Committee (No 78/01.04.2020). This study was registered in ClinicalTrials.gov, with number NCT04705753.

## Results

### Protective effect of CAPeo on VERO cells

The potential antiviral effect of CAPeo against SARS-CoV-2 was assessed both at cellular and molecular level, *in vitro*. Cells infected with SARS-CoV-2 present a CPE effect. CPE of infected VERO cells, was monitored in the presence of different concentrations of CAPeo (Figure 1A). As shown, cells were viable and presented a morphology similar to that of non-infected cells, up to a concentration of 1/100 CAPeo, where minimal CPE was present. At lower concentrations, CPE was obvious and cell morphology was more similar to the vehicle (DMSO)-treated cells. Similar results were found in cells preincubated with the same concentrations of CAPeo, for 2h, before infection with SARS-CoV-2 (0.1 m.o.i.).

**Figure 1.**
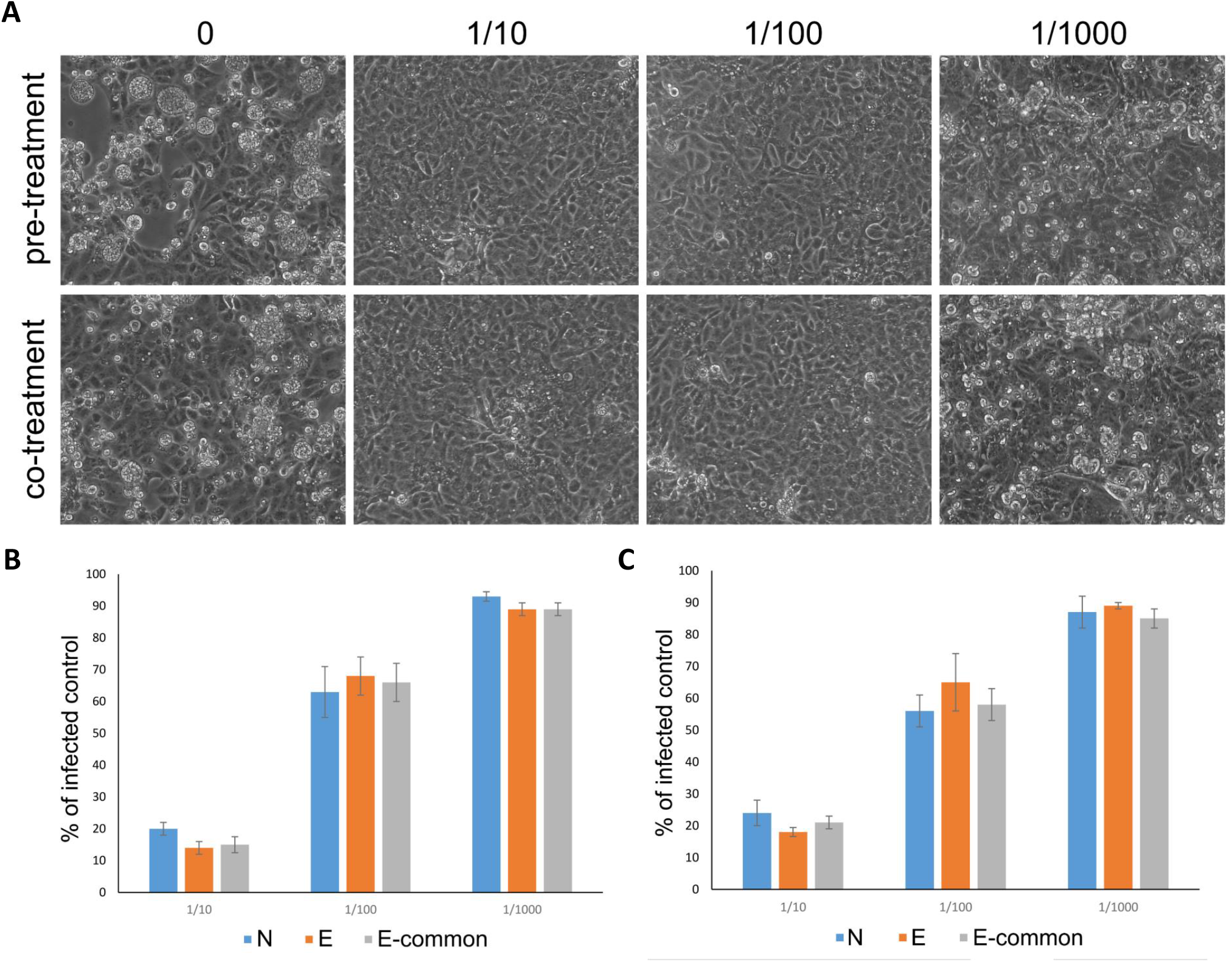
*In vitro* effect of CAPeo on SARS-Cov2 induced CPE and SARS-Cov2 replication. **A**. Light microscopy photographs of CPE in control (0, DMSO) and SARS-CoV-2 infected VERO cells (0.1 m.o.i), pre-treated or co-treated with different concentrations of CAPeo, in DMSO. **B**. Bar chart representing relative abundance (% of untreated control) of SARS-Cov2 RNA after pre-treatment (B) or co-treatment (C) with different concentrations of CAPeo, using real-time quantitative RT-PCR, targeting N and E regions of SARS-Cov2 genome and E-common region shared by SARS-CoV and SARS-CoV-2 viruses.

To avoid drug carry-over effects during TCID_50_ and to assess more accurately viral RNA production, we used quantitative real time PCR for the determination of viral RNA presence in culture medium. Real-time PCR analysis of three different SARS-CoV-2 genes (N, E, and SARS-CoV/SARS-CoV-2 common E region), from the supernatant of infected cells, treated with different concentrations of CAPeo, is shown in Figure 1B. Results (expressed as % of non-treated cells) showed that at a concentration of 1/10, CAPeo significantly reduced the viral release to the medium by >80%. The effect, albeit smaller (∼35%), persisted at concentrations of CAPeo 1/100 of the suggested dose in humans, but is absent at concentrations 1/1000. The calculated IC_50_ of CAPeo, with a logistic curve fitting, is 1/60 of the proposed dose for viral growth and ∼1/250 for maintenance of the cell phenotype. Interestingly, similar results (Figure 1C) were found when VERO cells were preincubated with different concentrations of CAPeo for 2h prior infection, suggesting that CAPeo, in addition to a possible therapeutic action, might be dotted with a prophylactic effect against SARS-CoV-2 virus.

### Effect of a 14-days CAPeo administration in humans on the severity and duration of symptoms of COVID-19-positive individuals. A proof-of-concept study

Results, presented above, suggest a direct SARS-CoV-2 inhibitory effect of CAPeo in cells. In order to provide a proof of concept on the effect of the preparation in humans, we have performed a small intervention study in 17 COVID-19-positive individuals, with mild symptoms, not necessitating hospitalization, in the context of a primary care center. No adverse effects were noted in any patient, in accordance with our previous observations in humans (Duijker et al., 2015;Anastasaki et al., 2017) and experimental animals (Kalyvianaki et al., 2020). The average age of enrolled patients was 34.4±11.0 years and underlying morbidities were reported in 29.4% of them; co-morbidities included dyslipidaemia, obesity, COPD and Hashimoto disease. Based on the questionnaire used, infection source was established to be either cohabitation (23.5%) or close contact with a confirmed case (58.8%).

On day 1, these individuals have consulted their General Practitioner (GP), with a variety of symptoms, both general and local (Figure 2A). Interestingly, general symptoms (fever, which in our population was mild, lower than 37.5°C, in all that two individuals, headache, fatigue and myalgias) were similar to those previously reported in two of the largest studies, focusing on symptom duration for COVID-19 outpatients in the US (Tenforde et al., 2020) and on the clinical presentation of mild-to-moderate forms of COVID-19 in Europe (Lechien et al., 2020). However, in our group, the frequency of nasal congestion, cough, anosmia and ageusia was lower.

**Figure 2.**
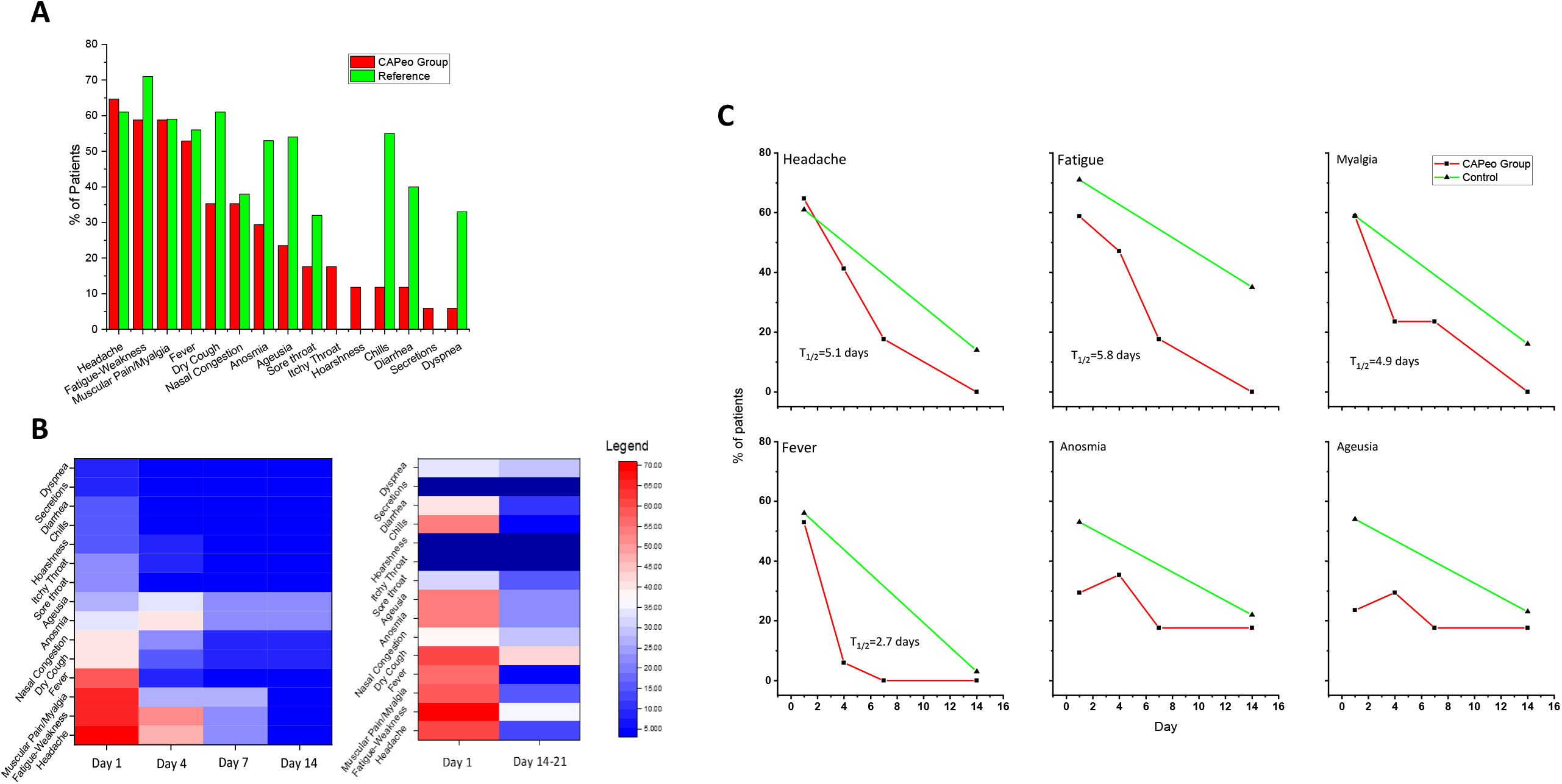
CAPeo administration in humans ameliorates symptoms of mild COVID-19 in ambulatory patients. **A**. Frequency of symptoms, common to both studies at Day 1, in our group and in (Tenforde et al., 2020). **B**. Heatmaps of symptom frequency in our CAPeo-treated group (left panel) and the population reported by (Tenforde et al., 2020) (right panel). **C**. Evolution of selected symptoms in our CAPeo-treated group (red curves). T_1/2_ for the resolution of symptoms was calculated with a logistic regression fit, with Origin Pro 2018. For comparison, the frequency of symptoms in the reference population reported by (Tenforde et al., 2020) is also presented (green curves). In panels B-C, the frequency of symptoms was extracted from Figure 1 of (Tenforde et al., 2020), with the online resource WebPlotDigitizer (Rohatgi, 2020).

In our study group, we have observed that the number of symptoms (4 at day1, 2 at day 4 and 1 at day 7) and the frequency of symptoms significantly decreased after CAPeo administration (Table 2, Figure 2B, left panel). The severity of symptoms, measured as the sum of symptoms in the seven-point Likert scale (see Material and Methods) was also decreased, following CAPeo administration (Wilcoxon tests; p<0.01, Table 2). Interestingly, at day 14, almost all symptoms completely regressed, with the notable exception of anosmia and ageusia, and mild ENT symptoms (nasal congestion and cough, in a small number of patients). EC_20_, EC_50_ and EC_80_, calculated with a logistic fit from data shown in Table 2 for the totality of symptom frequency and intensity was 2.3, 3.7 and 5.7 days respectively. Unfortunately, there are not many studies reporting the evolution of COVID-19 symptoms for comparison. From the two large published studies (Allen et al., 2020;Tenforde et al., 2020), we have extracted values from reported figures, with the help of the web resource WebPlotDigitizer (https://apps.automeris.io/wpd) (Rohatgi, 2020). Data from (Tenforde et al., 2020) are reported in Figure 2B, right panel. Fourteen days after COVID-19 testing, a number of symptoms persist; they include headache, fatigue, myalgias, anosmia, ageusia and respiratory distress. Concerning the common general symptoms (Figure 2C), we observe a notable, significant difference in the frequency of occurrence of headache, fatigue and myalgias, while fever was absent in both groups and a similar proportion of patients with persisting anosmia and ageusia.

**Table 2.** Change of total number of symptoms recorded on Day 1, 4, 7 and 14.

|  |  | Consultation Day Recording |  |  |  |  |
| --- | --- | --- | --- | --- | --- | --- |
|  |  | 1 | 4 | 7 | 14 |  |
| Number of symptoms |  | 75 | 38 | 18 | 8 | p-value |
| Δ-change (%) | 1st → 4th | | -37 (-49.3%) | | | 0.009 |
|  | 1st → 7th |  |  | -57 (-76.0%) |  | <0.001 |
|  | 1st → 14th |  |  |  | -67 (-89.3%) | <0.001 |
|  | 4th → 7th |  |  | -20 (-52.6%) |  | 0.004 |
| Wilcoxon tests (Monte Carlo simulation) |  |  |  |  |  |  |

Symptoms evolution was compared to that reported by (Tenforde et al., 2020); the authors report symptoms at days 1 and 14. In another study, we have used also as reference (Allen et al., 2020), including a much larger number of cases, symptoms were self-reported, daily, but without the implication of a medical examiner. We also used these data for comparison, as the frequency of symptoms was significantly different from the two other studies (Lechien et al., 2020;Tenforde et al., 2020), at day 1. A complete resolution of headache, a major symptom in COVID-19, was found in our group, with T_1/2_ of 5.1 days, while it persisted in 14% of cases in (Tenforde et al., 2020) and was almost absent in (Allen et al., 2020). Fatigue was also completely resolved in our group with T_1/2_ of 5.8 days, while it persisted in 35% of patients in the study of (Tenforde et al., 2020) and at about 15% in (Allen et al., 2020). Fever, another major symptom in day 1, completely resolved in our group, with T_1/2_ of 2.7 days, as compared to ∼9 days in (Allen et al., 2020). At 14 days, fever was also absent in studies by (Allen et al., 2020;Tenforde et al., 2020). Therefore, CAPeo seems to ameliorate general symptoms very quickly, better than the reference population. In addition, as shown in Figure 2B, the majority of symptoms, both general and local, completely resolve at the end of the first week. A special notion applies to anosmia and ageusia. Both in our group and in the European multi-center study (Allen et al., 2020), these symptoms progressively increase, peaking at day 4 and at days 4-6 respectively. Thereafter, at day 14, these two symptoms persist in 17%, 23% and 21% in our group, in (Tenforde et al., 2020) and in (Allen et al., 2020) studies, respectively.

As mentioned, and according to protocol, we included three subjects in the analysis since they were family members (father, mother and sister of one patient), working together and living in the same house. They received the CAPeo for prophylactic use, on the basis of the decision of the family doctor, despite having tested negative for SARS-CoV-2. One of them (mother) presented symptoms two days after the baseline, and was re-tested and found positive, and she was enrolled in the study, while the remaining two (father and sister) did not report symptoms on any of the days of observation. We consider this an important reporting aspect in terms of establishing guidelines for sequential testing, managing oligo- or asymptomatic patients, in outpatient settings and to inform future clinical study design across settings, as highlighted by a recent report (Wernhart et al., 2020). The median incubation period of five days creates a false sense of safety, but also presents a challenge in terms of study inclusion and sound trial conduct and reporting (Lauer et al., 2020).

## Discussion

The COVID-19 pandemics imposed a number of, not yet resolved, problems to the International Scientific Community. Thanks to the combined world-wide scientific effort and the analysis of SARS-CoV-2 virus (O’Leary and Ovsepian, 2020), successful and safe vaccines are now begin to emerge (Polack et al., 2020;Voysey et al., 2020). Although multiple molecules are at various stages of preclinical and clinical development, there largely remains an unmet need for prophylactic and therapeutic regimens to combat the disease, with proposed measures being scarce and non-specific (Bolarin et al., 2020;Wang et al., 2020a), with the exception of monoclonal antibodies (Baum et al., 2020;Hansen et al., 2020;Weinreich et al., 2020) and dexamethasone (Cain and Cidlowski, 2020;Recovery Collaborative Group et al., 2020), which primarily target hospitalized patients in intensive care units. In this respect, the need for non-expensive therapeutic regimens, safe and effective in non-critically-ill patients and efficient for their management in ambulatory settings, remain unmet. Accordingly, drug repurposing, for candidates acting against known or predicted SARS-CoV-2 protein actions have been advanced (reviewed and discussed in (Asselah et al., 2020;Cadegiani, 2020;Khan et al., 2020)), while natural products have also been tested (reviewed in (Benarba and Pandiella, 2020)). Finally, quercetin has been proposed as an alternative for dexamethasone (Pawar and Pal, 2020). Here, we suggest CAPeo as a potential novel agent, for the safe and effective therapeutic management of ambulatory mild cases of COVID-19.

CAPeo, a 1.5% of essential oils of *Thymbra capitata* (L.) Cav., *Salvia fruticosa* Mill. and *Origanum dictamnus* L. in olive oil, has been advanced by our group in 2015, and found to be effective in reducing the severity and duration of symptoms of viral upper respiratory tract infections (Duijker et al., 2015;Anastasaki et al., 2017). It presents remarkable anti-viral properties against Influenza A and B strains and HRV14 (Tseliou et al., 2019), while it is safe in both experimental animals (Kalyvianaki et al., 2020) and humans (Duijker et al., 2015). Its properties have been recently reviewed in reference (Pirintsos et al., 2020). Here, we extend these previous findings, by providing *in vitro* evidence about its antiviral activity against SARS-CoV-2 infected VERO cells. CAPeo was effective at concentrations compatible with the expected circulating concentrations of CAPeo constituents (Scalbert et al., 2002;Manach et al., 2005), and similar with the previously reported *in vitro* antiviral activity in Influenza strains and HRV14 (Tseliou et al., 2019). Interestingly, as shown in Figure 1, CAPeo mixture was both prophylactic and therapeutic *in vitro*, at concentrations up to 1/100 the suggested *per os* dose in humans, preserving the viability, cell phenotype and viral RNA presence in the culture medium. In this respect, calculated EC_50_ through a logistic fit was estimated as 1/60 of the proposed dose for viral growth, and ∼1/250 for cell phenotype, compatible with the expected concentrations of CAPeo constituents in human plasma (Scalbert et al., 2002;Manach et al., 2005).

CAPeo contains 25 different micro-constituents (please refer to Supplemental Table 4 of Reference (Duijker et al., 2015), for an exhaustive presentation of concentrations of specific constituents). The main compounds are carvacrol (53%), eucalyptol (13%), β-Caryophyllene (3%), p-Cymene (1.32%), γ-Terpinene (1.17%), Borneol (1.68%) and α-Terpineol (1.06%). As reviewed recently (Pirintsos et al., 2020), none of these compounds have been reported as anti-virals. However, work in progress in our group has identified specific viral targets for some of these constituents, with a direct impact on viral replication.

Based on the encouraging *in vitro* results, and having ensured the safety of CAPeo in experimental animals (Kalyvianaki et al., 2020) and humans (Duijker et al., 2015), we have further performed a Proof-of-Concept intervention study in humans. Due to the very low incidence of COVID-19 positive cases in Crete, at the time of the study, only seventeen (17) eligible ambulatory patients, positive for COVID-19 by real-time quantitative PCR, were enrolled, and tested for the severity and duration of general and local symptoms, for 14 days. We have chosen this interval as previous studies report a self-resolution of mild COVID-19 cases in 14 (Tenforde et al., 2020), or 14-21 days (Allen et al., 2020) and the persistence of virus in upper respiratory tract samples for about 10 days (Singanayagam et al., 2020). A concrete limitation of this study is that we do not have a control group; we have therefore compared the evolution of disease symptoms with the few studies reporting the evolution of symptoms in non-hospitalized patients (Allen et al., 2020;Tenforde et al., 2020). As discussed previously (Allen et al., 2020;Lechien et al., 2020), symptoms may vary significantly, related to the ethnicity of participants. At the beginning of the study (patient consultation and positive real-time PCR result), the main general symptoms include headache, myalgia, weakness and fever, in accord with previous investigations (Allen et al., 2020;Lechien et al., 2020;Tenforde et al., 2020). However, in our group, fever was low <37.5°C in all but one patient, and the frequency and severity of other symptoms, such as gastrointestinal (diarrhea, respiratory and ENT symptoms were low, possibly because of the low viral load, but without excluding the possibility of simply witnessing an effect prevalent simply because of the small group size.

Symptoms evolution was compared to that reported symptoms at days 1 and 14, by (Tenforde et al., 2020) and (Allen et al., 2020), including a larger number of cases, in which symptoms were self-reported, daily, but without the participation of a medical examiner. In our group, treated with CAPeo, we report a complete resolution of headache, fatigue and fever, major general symptoms in COVID-19. In contrast, headache, and fatigue persisted in the studies of (Allen et al., 2020;Tenforde et al., 2020), while fever was equally resolved. We have therefore concluded that CAPeo ameliorates general symptoms very quickly, better than an untreated population. In addition, as shown in Figure 2B, the majority of symptoms, both general and local, almost completely resolve at the end of the first week. Although we have not analyzed in depth the underlying mechanism of action for this beneficial effect of CAPeo in the evolution of COVID-19, in addition to the possible direct anti-viral effect reported in cells, another mechanism of action might be its anti-inflammatory effect, previously reported in experimental animals (Kalyvianaki et al., 2020) and humans (Duijker et al., 2015). Anosmia and ageusia, however, evolved both in our group and in (Allen et al., 2020), presenting a maximum at day 4 and at days 4-6 respectively, and persist in 17%, 23% and 21% in our group, in the study of (Tenforde et al., 2020) and in the population reported by (Allen et al., 2020) respectively. Whether this is due to a late recovery of nasal and buccal mucosa, or in the persistence of virus in a small percentage of patients (Singanayagam et al., 2020) is not clear yet.

In conclusion, our findings suggest that CAPeo, a mixture of essential oils of three Cretan aromatic plants, possesses a potent antiviral activity, in addition to Influenza and HRV14 (Tseliou et al., 2019), against SARS-CoV-2, in which it also possesses a prophylactic activity. In addition, our reported here proof-of-concept intervention study in humans shows that it significantly reduces general and local symptoms of mild COVID-19 patients. If these results will be confirmed in a planned prospective clinical study, CAPeo might be a novel, inexpensive, therapeutic agent in cases of ambulatory COVID-19 patients.

## Data Availability

All data are included in the manuscript

## Author Contribution

CL conceived the Proof-of-Concept clinical study. GS, IK and EC designed the experimental studies. CL, EP, ES, and ML equally participated in the design of the Proof-of-Concept clinical study and the analysis thereof. EP, AD, and CL were responsible for trial conduct and all clinical operation aspects, including the preliminary trial report, whereas ML and EC performed the statistical analysis. GS performed the COVID-19 real-time quantitative PCR in clinical samples. SAP and MK participating in the design and analysis of experimental data. IK and MP performed the *in vitro* studies and IK drafted the initial report. EC wrote the first draft of the paper, while all authors contributed to the reduction of the manuscript. All authors approved the submission.

## References

Allen, W.E., Altae-Tran, H., Briggs, J., Jin, X., Mcgee, G., Shi, A., Raghavan, R., Kamariza, M., Nova, N., Pereta, A., Danford, C., Kamel, A., Gothe, P., Milam, E., Aurambault, J., Primke, T., Li, W., Inkenbrandt, J., Huynh, T., Chen, E., Lee, C., Croatto, M., Bentley, H., Lu, W., Murray, R., Travassos, M., Coull, B.A., Openshaw, J., Greene, C.S., Shalem, O., King, G., Probasco, R., Cheng, D.R., Silbermann, B., Zhang, F., and Lin, X. (2020). Population-scale longitudinal mapping of COVID-19 symptoms, behaviour and testing. Nat Hum Behav 4, 972–982.

Anastasaki, M., Bertsias, A., Pirintsos, S.A., Castanas, E., and Lionis, C. (2017). Post-market outcome of an extract of traditional Cretan herbs on upper respiratory tract infections: a pragmatic, prospective observational study. BMC Complement Altern Med 17, 466.

Asselah, T., Durantel, D., Pasmant, E., Lau, G., and Schinazi, R.F. (2020). COVID-19: discovery, diagnostics and drug development. J Hepatol.

Bariotakis, M., Georgescu, L., Laina, D., Oikonomou, I., Ntagounakis, G., Koufaki, M.I., Souma, M., Choreftakis, M., Zormpa, O.G., Smykal, P., Sourvinos, G., Lionis, C., Castanas, E., Karousou, R., and Pirintsos, S.A. (2019). From wild harvest towards precision agriculture: Use of Ecological Niche Modelling to direct potential cultivation of wild medicinal plants in Crete. Sci Total Environ 694, 133681.

Baum, A., Ajithdoss, D., Copin, R., Zhou, A., Lanza, K., Negron, N., Ni, M., Wei, Y., Mohammadi, K., Musser, B., Atwal, G.S., Oyejide, A., Goez-Gazi, Y., Dutton, J., Clemmons, E., Staples, H.M., Bartley, C., Klaffke, B., Alfson, K., Gazi, M., Gonzalez, O., Dick, E., Jr., Carrion, R., Jr., Pessaint, L., Porto, M., Cook, A., Brown, R., Ali, V., Greenhouse, J., Taylor, T., Andersen, H., Lewis, M.G., Stahl, N., Murphy, A.J., Yancopoulos, G.D., and Kyratsous, C.A. (2020). REGN-COV2 antibodies prevent and treat SARS-CoV-2 infection in rhesus macaques and hamsters. Science.

Benarba, B., and Pandiella, A. (2020). Medicinal Plants as Sources of Active Molecules Against COVID-19. Front Pharmacol 11, 1189.

Bolarin, J.A., Oluwatoyosi, M.A., Orege, J.I., Ayeni, E.A., Ibrahim, Y.A., Adeyemi, S.B., Tiamiyu, B.B., Gbadegesin, L.A., Akinyemi, T.O., Odoh, C.K., Umeobi, H.I., and Adeoye, A.B. (2020). Therapeutic drugs for SARS-CoV-2 treatment: Current state and perspective. Int Immunopharmacol 90, 107228.

Cadegiani, F.A. (2020). Repurposing existing drugs for COVID-19: an endocrinology perspective. BMC Endocr Disord 20, 149.

Cain, D.W., and Cidlowski, J.A. (2020). After 62 years of regulating immunity, dexamethasone meets COVID-19. Nat Rev Immunol 20, 587–588.

Duijker, G., Bertsias, A., Symvoulakis, E.K., Moschandreas, J., Malliaraki, N., Derdas, S.P., Tsikalas, G.K., Katerinopoulos, H.E., Pirintsos, S.A., Sourvinos, G., Castanas, E., and Lionis, C. (2015). Reporting effectiveness of an extract of three traditional Cretan herbs on upper respiratory tract infection: results from a double-blind randomized controlled trial. J Ethnopharmacol 163, 157–166.

Hansen, J., Baum, A., Pascal, K.E., Russo, V., Giordano, S., Wloga, E., Fulton, B.O., Yan, Y., Koon, K., Patel, K., Chung, K.M., Hermann, A., Ullman, E., Cruz, J., Rafique, A., Huang, T., Fairhurst, J., Libertiny, C., Malbec, M., Lee, W.Y., Welsh, R., Farr, G., Pennington, S., Deshpande, D., Cheng, J., Watty, A., Bouffard, P., Babb, R., Levenkova, N., Chen, C., Zhang, B., Romero Hernandez, A., Saotome, K., Zhou, Y., Franklin, M., Sivapalasingam, S., Lye, D.C., Weston, S., Logue, J., Haupt, R., Frieman, M., Chen, G., Olson, W., Murphy, A.J., Stahl, N., Yancopoulos, G.D., and Kyratsous, C.A. (2020). Studies in humanized mice and convalescent humans yield a SARS-CoV-2 antibody cocktail. Science 369, 1010–1014.

Kalyvianaki, K., Malamos, P., Mastrodimou, N., Manoura-Zonou, I., Vamvoukaki, R., Notas, G., Malliaraki, N., Moustou, E., Tzardi, M., Pirintsos, S., Lionis, C., Sourvinos, G., Castanas, E., and Kampa, M. (2020). Toxicity evaluation of an essential oil mixture from the Cretan herbs thyme, Greek sage and Cretan dittany. NPJ Sci Food 4, 20.

Khan, Z., Ghafoor, D., Khan, A., Ualiyeva, D., Khan, S.A., Bilal, H., Khan, B., Khan, A., and Sajjad, W. (2020). Diagnostic approaches and potential therapeutic options for coronavirus disease (COVID-19). New Microbes New Infect, 100770.

Lauer, S.A., Grantz, K.H., Bi, Q., Jones, F.K., Zheng, Q., Meredith, H.R., Azman, A.S., Reich, N.G., and Lessler, J. (2020). The Incubation Period of Coronavirus Disease 2019 (COVID-19) From Publicly Reported Confirmed Cases: Estimation and Application. Ann Intern Med 172, 577–582.

Lechien, J.R., Chiesa-Estomba, C.M., Place, S., Van Laethem, Y., Cabaraux, P., Mat, Q., Huet, K., Plzak, J., Horoi, M., Hans, S., Rosaria Barillari, M., Cammaroto, G., Fakhry, N., Martiny, D., Ayad, T., Jouffe, L., Hopkins, C., Saussez, S., and Yo-Ifos, C.-T.F.O. (2020). Clinical and epidemiological characteristics of 1420 European patients with mild-to-moderate coronavirus disease 2019. J Intern Med 288, 335–344.

Manach, C., Williamson, G., Morand, C., Scalbert, A., and Remesy, C. (2005). Bioavailability and bioefficacy of polyphenols in humans. I. Review of 97 bioavailability studies. Am J Clin Nutr 81, 230S–242S.

O’leary, V.B., and Ovsepian, S.V. (2020). Severe Acute Respiratory Syndrome Coronavirus 2 (SARS-CoV-2). Trends Genet 36, 892–893.

Pawar, A., and Pal, A. (2020). Molecular and functional resemblance of dexamethasone and quercetin: A paradigm worth exploring in dexamethasone-nonresponsive COVID-19 patients. Phytother Res.

Pirintsos, S.A., Bariotakis, M., Kampa, M., Sourvinos, G., Lionis, C., and Castanas, E. (2020). The Therapeutic Potential of the Essential Oil of Thymbra capitata (L.) Cav., Origanum dictamnus L. and Salvia fruticosa Mill. And a Case of Plant-Based Pharmaceutical Development. Frontiers in Pharmacology 11, 522213.

Polack, F.P., Thomas, S.J., Kitchin, N., Absalon, J., Gurtman, A., Lockhart, S., Perez, J.L., Perez Marc, G., Moreira, E.D., Zerbini, C., Bailey, R., Swanson, K.A., Roychoudhury, S., Koury, K., Li, P., Kalina, W.V., Cooper, D., Frenck, R.W., Jr., Hammitt, L.L., Tureci, O., Nell, H., Schaefer, A., Unal, S., Tresnan, D.B., Mather, S., Dormitzer, P.R., Sahin, U., Jansen, K.U., Gruber, W.C., and Group, C.C.T. (2020). Safety and Efficacy of the BNT162b2 mRNA Covid-19 Vaccine. N Engl J Med.

Recovery Collaborative Group, Horby, P., Lim, W.S., Emberson, J.R., Mafham, M., Bell, J.L., Linsell, L., Staplin, N., Brightling, C., Ustianowski, A., Elmahi, E., Prudon, B., Green, C., Felton, T., Chadwick, D., Rege, K., Fegan, C., Chappell, L.C., Faust, S.N., Jaki, T., Jeffery, K., Montgomery, A., Rowan, K., Juszczak, E., Baillie, J.K., Haynes, R., and Landray, M.J. (2020). Dexamethasone in Hospitalized Patients with Covid-19 - Preliminary Report. N Engl J Med.

Reed, L.J., and Muench, H. (1938). A simple method of estimating fifty per cent endpoints. Am. J. Epidemiol. 27, 493–497.

Rohatgi, A. (2020). WebPlotDigitizer [Online]. Pacifica, California, USA: Automeris.io. Available: https://automeris.io/WebPlotDigitizer [Accessed].

Scalbert, A., Morand, C., Manach, C., and Remesy, C. (2002). Absorption and metabolism of polyphenols in the gut and impact on health. Biomed Pharmacother 56, 276–282.

Singanayagam, A., Patel, M., Charlett, A., Lopez Bernal, J., Saliba, V., Ellis, J., Ladhani, S., Zambon, M., and Gopal, R. (2020). Duration of infectiousness and correlation with RT-PCR cycle threshold values in cases of COVID-19, England, January to May 2020. Euro Surveill 25.

Tenforde, M.W., Kim, S.S., Lindsell, C.J., Billig Rose, E., Shapiro, N.I., Files, D.C., Gibbs, K.W., Erickson, H.L., Steingrub, J.S., Smithline, H.A., Gong, M.N., Aboodi, M.S., Exline, M.C., Henning, D.J., Wilson, J.G., Khan, A., Qadir, N., Brown, S.M., Peltan, I.D., Rice, T.W., Hager, D.N., Ginde, A.A., Stubblefield, W.B., Patel, M.M., Self, W.H., Feldstein, L.R., Investigators, I.V.Y.N., Team, C.C.-R., and Investigators, I.V.Y.N. (2020). Symptom Duration and Risk Factors for Delayed Return to Usual Health Among Outpatients with COVID-19 in a Multistate Health Care Systems Network - United States, March-June 2020. MMWR Morb Mortal Wkly Rep 69, 993–998.

Tseliou, M., Pirintsos, S.A., Lionis, C., Castanas, E., and Sourvinos, G. (2019). Antiviral effect of an essential oil combination derived from three aromatic plants (Coridothymus capitatus (L.) Rchb. f., Origanum dictamnus L. and Salvia fruticosa Mill.) against viruses causing infections of the upper respiratory tract. Journal of Herbal Medicine 17–18.

Voysey, M., Clemens, S.a.C., Madhi, S.A., Weckx, L.Y., Folegatti, P.M., Aley, P.K., Angus, B., Baillie, V.L., Barnabas, S.L., Bhorat, Q.E., Bibi, S., Briner, C., Cicconi, P., Collins, A.M., Colin-Jones, R., Cutland, C.L., Darton, T.C., Dheda, K., Duncan, C.J.A., Emary, K.R.W., Ewer, K.J., Fairlie, L., Faust, S.N., Feng, S., Ferreira, D.M., Finn, A., Goodman, A.L., Green, C.M., Green, C.A., Heath, P.T., Hill, C., Hill, H., Hirsch, I., Hodgson, S.H.C., Izu, A., Jackson, S., Jenkin, D., Joe, C.C.D., Kerridge, S., Koen, A., Kwatra, G., Lazarus, R., Lawrie, A.M., Lelliott, A., Libri, V., Lillie, P.J., Mallory, R., Mendes, A.V.A., Milan, E.P., Minassian, A.M., Mcgregor, A., Morrison, H., Mujadidi, Y.F., Nana, A., O’reilly, P.J., Padayachee, S.D., Pittella, A., Plested, E., Pollock, K.M., Ramasamy, M.N., Rhead, S., Schwarzbold, A.V., Singh, N., Smith, A., Song, R., Snape, M.D., Sprinz, E., Sutherland, R.K., Tarrant, R., Thomson, E.C., Torok, M.E., Toshner, M., Turner, D.P.J., Vekemans, J., Villafana, T.L., Watson, M.E.E., Williams, C.J., Douglas, A.D., Hill, A.V.S., Lambe, T., Gilbert, S.C., Pollard, A.J., and Oxford, C.V.T.G. (2020). Safety and efficacy of the ChAdOx1 nCoV-19 vaccine (AZD1222) against SARS-CoV-2: an interim analysis of four randomised controlled trials in Brazil, South Africa, and the UK. Lancet.

Wang, M.Y., Zhao, R., Gao, L.J., Gao, X.F., Wang, D.P., and Cao, J.M. (2020a). SARS-CoV-2: Structure, Biology, and Structure-Based Therapeutics Development. Front Cell Infect Microbiol 10, 587269.

Wang, Y., Huo, P., Dai, R., Lv, X., Yuan, S., Zhang, Y., Guo, Y., Li, R., Yu, Q., and Zhu, K. (2020b). Convalescent plasma may be a possible treatment for COVID-19: A systematic review. Int Immunopharmacol 91, 107262.

Weinreich, D.M., Sivapalasingam, S., Norton, T., Ali, S., Gao, H., Bhore, R., Musser, B.J., Soo, Y., Rofail, D., Im, J., Perry, C., Pan, C., Hosain, R., Mahmood, A., Davis, J.D., Turner, K.C., Hooper, A.T., Hamilton, J.D., Baum, A., Kyratsous, C.A., Kim, Y., Cook, A., Kampman, W., Kohli, A., Sachdeva, Y., Graber, X., Kowal, B., Dicioccio, T., Stahl, N., Lipsich, L., Braunstein, N., Herman, G., Yancopoulos, G.D., and Trial, I. (2020). REGN-COV2, a Neutralizing Antibody Cocktail, in Outpatients with Covid-19. N Engl J Med.

Wernhart, S., Forster, T.H., and Weihe, E. (2020). Outpatient Management of Oligosymptomatic Patients with respiratory infection in the era of SARS-CoV-2: Experience from rural German general practitioners. BMC Infect Dis 20, 811.

